# COVID-19 infection results in alterations of the kynurenine pathway and fatty acid metabolism that correlate with IL-6 levels and renal status

**DOI:** 10.1101/2020.05.14.20102491

**Authors:** Tiffany Thomas, Davide Stefanoni, Julie A. Reisz, Travis Nemkov, Lorenzo Bertolone, Richard O Francis, Krystalyn E. Hudson, James C. Zimring, Kirk C. Hansen, Eldad A. Hod, Steven L. Spitalnik, Angelo D’Alessandro

**Author notes:** Equally contributing last/senior authors (Steven L. Spitalnik, Angelo D’Alessandro). Corresponding authors: Angelo D’Alessandro, PhD Department of Biochemistry and Molecular Genetics University of Colorado Anschutz Medical Campus 12801 East 17th Ave., Aurora, CO 80045 Steven L. Spitalnik, MD Department of Pathology & Cell Biology Columbia University 630 West 168th Street New York, NY 10032.

## Abstract

Previous studies suggest a role for systemic reprogramming of host metabolism during viral pathogenesis to fuel rapidly expanding viral proliferation, for example by providing free amino acids and fatty acids as building blocks. In addition, general alterations in metabolism can provide key understanding of pathogenesis. However, little is known about the specific metabolic effects of SARS-COV-2 infection. The present study evaluated the serum metabolism of COVID-19 patients (n=33), identified by a positive nucleic acid test of a nasopharyngeal swab, as compared to COVID-19-negative control patients (n=16). Targeted and untargeted metabolomics analyses specifically identified alterations in the metabolism of tryptophan into the kynurenine pathway, which is well-known to be involved in regulating inflammation and immunity. Indeed, the observed changes in tryptophan metabolism correlated with serum interleukin-6 (IL-6) levels. Metabolomics analysis also confirmed widespread dysregulation of nitrogen metabolism in infected patients, with decreased circulating levels of most amino acids, except for tryptophan metabolites in the kynurenine pathway, and increased markers of oxidant stress (e.g., methionine sulfoxide, cystine), proteolysis, and kidney dysfunction (e.g., creatine, creatinine, polyamines). Increased circulating levels of glucose and free fatty acids were also observed, consistent with altered carbon homeostasis in COVID-19 patients. Metabolite levels in these pathways correlated with clinical laboratory markers of inflammation and disease severity (i.e., IL-6 and C-reactive protein) and renal function (i.e., blood urea nitrogen). In conclusion, this initial observational study of the metabolic consequences of COVID-19 infection in a clinical cohort identified amino acid metabolism (especially kynurenine and cysteine/taurine) and fatty acid metabolism as correlates of COVID-19, providing mechanistic insights, potential markers of clinical severity, and potential therapeutic targets.

**Key points:**

- Metabolism of tryptophan into the kynurenine pathway is increased in COVID-19 patients
- COVID-19 infection is significantly associated with dysregulated nitrogen and carbon metabolism
- COVID-19 infection induces increased circulating levels of free fatty acids and glucose

## Introduction

On December 12, 2019 the first patient presented in Wuhan, Hubei province, China with symptoms consistent with severe acute respiratory syndrome (SARS). Soon investigators realized that these symptoms were caused by a new viral infection, as gleaned by a metagenomics analysis of the bronchoalveolar lavage fluid from this patient. The newly identified RNA virus strain belongs to the *Coronaviridae* family and was referred to as ‘2019-nCoV’.(1) Phylogenetic analyses of the genome (29,903 nucleotides) demonstrated its similarity (89.1%) to a group of SARS-like coronaviruses (genus *Betacoronavirus*, subgenus *Sarbecovirus*) that were previously isolated in bats in China (1). Since then, the virus spread around the world, resulting in 2,959,929 confirmed cases and 202,733 confirmed deaths as of April 28, 2020 (https://www.who.int/emergencies/diseases/novel-coronavirus-2019).

To face the global pandemic, entire nations initiated lockdowns, social distancing, economic peril, and mandatory regulations regarding wearing personal protective equipment in public. In parallel, a worldwide mobilization of scientists has occurred, in an effort to slow viral spread, identify suitable therapies for infected patients, and expedite vaccine development and testing (2). This resulted in the rapid generation of new information, as well as a concomitant concern for the quality of the evidence being disseminated (3).

COVID-19 initiates a wide spectrum of symptoms, including fever, persistent dry cough, shortness of breath, chills, muscle pain, headache, loss of taste or smell, and gastrointestinal symptoms. However, some subjects are more susceptible, by unknown mechanisms, to develop severe disease, especially older males with various comorbidities, including obesity, diabetes, cardiovascular disease, or immunosuppression (e.g., cancer patients undergoing chemo- or radio-therapy, or transplant patients). In contrast, children, younger adults, and women tend to be either asymptomatic or present with mild disease, while still being contagious and contributing to viral transmission. As such, widespread testing, even of asymptomatic subjects, has been advocated as a strategy to monitor and prevent disease dissemination. Testing nasopharyngeal swabs with nucleic acid amplification assays was introduced worldwide to diagnose disease and monitor recovery; for example, Italy, one of the countries hit hardest by COVID-19, enforces quarantine of any positive subjects until they no longer show symptoms for at least 14 days and test negative twice within 24h. However, logistical issues have limited generalized testing in most countries, including the United States (<12,000 tests/million people) (4). Nonetheless, recent development of serological testing for antibodies against SARS-CoV-2 proteins suggest that a significantly larger fraction of the total population may have been infected than that identified by testing nasopharyngeal swabs (e.g., up to 85-fold higher than molecularly-confirmed cases) (5). These findings provoked ongoing debates and controversy, especially regarding test sensitivity and specificity, and the distinction between case fatality rates (mortality among those who tested positive by molecular assays) and infection fatality rates (extrapolated from serological testing results), resulting in estimates of ∼75-fold and ∼7-fold higher mortality than seasonal influenza, respectively. Large international initiatives (e.g., Solidarity II) propose to tackle this issue through homogeneous serological testing in 12 countries (6). At the center of this debate, serological testing detects antibodies against SARS-COV-2 proteins in patients who have recovered from infection. These patients are now being actively enrolled globally as “convalescent” plasma donors to determine whether their antibodies can help stop the spread of the virus and provide therapeutic benefit in severely ill COVID-19 patients (7).

From a molecular standpoint, recombinant expression of SARS-COV-2 proteins, along with structural and protein-protein interaction (8) studies, confirm that, like other phylogenetically-close (9) coronaviruses, the SARS-COV-2 spike protein S interacts with the angiotensin converting enzyme receptor 2 (ACE2) to penetrate host cells, especially in the lung epithelium (10). Disruption of this interaction may provide a therapeutic target (11, 12); indeed, it is the strategy of several current vaccine candidates to elicit humoral responses to capsid proteins (2, 13). Still, it is unclear whether and for how long previously-positive recovered patients are immune to the virus and, therefore, the scientific community is invested in defining suitable interventions to treat the most severe symptoms of COVID-19. For example, in early March, 2020, preliminary evidence seemed to suggest a beneficial role for antimalarial drugs (e.g., hydroxycholoroquine), especially in combination with azithromycin, in decreasing viral load and improving the prognosis of COVID-19 patients (14); however, a larger clinical trial showed a deleterious effect (e.g., a ∼2-fold increase in mortality) for patients on hydroxycholoroquine (with or without azithromycin) (15). Other treatments, while totally effective in sanitizing inert surfaces (e.g., UV light or bleach) (16); are clearly incompatible with the “*primo non nocere*” principle of medical practice. Although using antiretroviral drugs to treat SARS coronaviruses was previously proposed *in vitro* (17), no clinical evidence *in vivo* has been provided to date; nonetheless, the preliminary release of data from ongoing trials with remdesivir show promise, with ∼30% faster recovery and reduction in mortality (ClinicalTrials.gov Identifier: NCT04280705).

Given the current difficulties in preventing infection or decreasing viral load, multiple approaches are being investigated to identify key pathways involved in the most serious sequelae of SARS-COV-2 infection. For example, a common finding in the most severe cases is “cytokine storm,” driven mostly by sustained increases in circulating levels of pro-inflammatory interleukin-6 (IL-6).(18) Antibody-based approaches (e.g., tocilizumab) are currently being tested as a strategy to mitigate the clinical complications of COVID-19-induced cytokine (18). Notably, this extreme inflammation often results in the need for ventilator support or, even, extracorporeal membrane oxygenation (19). However, in ∼80% of cases, the latter did not prevent mortality, owing to insufficient lung perfusion, which could be explained by the development of thromboembolic complications (20). In this context, clinical trials are currently underway to determine the impact of anticoagulants (e.g., heparin) or pro-fibrinolytic drugs (e.g., tissue plasminogen activator) in ameliorating severe COVID-19 infection with thromboembolic complications (21). Interestingly, platelets from older subjects are hypercoagulable in the presence of pro-inflammatory stimuli (22), which may help explain the increased mortality rates in older COVID-19 patients. Of note, small molecule metabolites are relevant in treating coagulopathies (e.g., tranexamic acid to treat hyperfibrinolysis in actively bleeding patients) (23). However, to date, nothing is known about the serum metabolome of COVID-19 patients.

Small molecule metabolites are essential for viral infection, because they provide building blocks that a rapidly proliferating virus requires to assemble its nucleic acids, proteins (including capsid proteins that mediate infection of new cell targets), and membrane. Indeed, viral infections mobilize free fatty acids to support viral capsid-associated membrane formation, which was described for other coronaviruses and is explained, in part, by activation of phospholipase A2, a target amenable to pharmacological intervention (24–26). The current study provides the first comprehensive targeted and untargeted metabolomics analysis of sera from COVID-19 patients, stratified by circulating levels of IL-6, and correlated to inflammatory markers and renal function. Though observational, this study identifies significant alterations in fatty acid metabolism in infected patients and unanticipated alterations of glucose homeostasis and amino acid metabolism; specifically, dysregulated tryptophan, arginine, and sulfur metabolism are potential therapeutic targets for COVID-19.

## Results

Forty-nine subjects were enrolled in this observational study: 33 COVID-19-positive subjects and 16 COVID-19-negative subjects, the latter included “never positive” subjects and convalescent plasma donors (**Figure 1.A**). Owing to the skewed demographics of the COVID-19-positive patients (prevalence and mortality being ∼fold higher in males) (27), 76% of the subjects in this group were male, 56.5 + 18.1 years old (mean + standard deviation) versus 38% male, 37.8 + 11.6 years old for the control group. Of the COVID-19-positive patients, 5 were asymptomatic or had mild symptoms with corresponding IL-6 levels of <10 pg/ml; 10 subjects had IL-6 levels of 10–65 pg/ml, and 18 subjects IL-6 levels of > 90 pg/ml. Subjects in these three sub-groups had no significant differences in age or sex. Reference ranges for IL-6 levels in healthy controls are <5 pg/ml (**Figure 1.B**). Of all the COVID-19-positive subjects, only 6 had renal function parameters within the reference range (blood urea nitrogen (BUN) of 7–26 mg/dl and creatinine of 0.5–0.95 mg/dl), of which 3 were in the low and 2 in the moderate IL-6 group, respectively (**Supplementary Table 1**). Targeted metabolomics analyses were performed on sera using UHPLC-MS and the results are reported extensively in **Supplementary Table 1**. The metabolic phenotypes of sera from COVID-19-positive patients differed significantly from controls, when examined by partial least-square discriminant analysis (PLS-DA; **Figure 1.C**). Hierarchical clustering analysis (**Figure 1.D**) highlighted significant associations of COVID-19 and IL-6 levels with amino acid metabolism, purines, acylcarnitines, and fatty acids; a vectorial version of this figure is provided in **Supplementary Figure 1**. Volcano plots (**Figure 1.E**) from targeted metabolomics analyses highlighted the top metabolites that increased (blue) or decreased (red) in the sera of COVID-19-positive patients, as compared to controls. To expand on these observations, untargeted metabolomics analyses were performed based on high-resolution, accurate intact mass, isotopic patterns and MS2 fragmentation patterns, as described;(28) these results are reported in **Supplementary Table 2**. In **Figure 2.A**, volcano plot analyses highlighted 3,034 and 2,484 differential metabolites for negative and positive ion modes between COVID-19-positive and –negative subjects, respectively. PLS-DA analysis based on untargeted metabolomics data confirmed the targeted results and further separated COVID-19-positive and COVID-19-negative subjects (**Figure 2.B**), the former group separating from the latter across principal component 1 (18.7% of the total variance) as a function of IL-6 levels. Interestingly, volcano plot analyses comparing COVID-19-positive subjects with low, medium, and high serum IL-6 levels, as compared to controls, showed that the low IL-6 group was most comparable to controls (199 and 218 metabolites significantly decreasing and increasing), whereas the medium (218 and 765) and high IL-6 (206 and 789) groups differed the most (**Figure 2.C**). Pathway analysis of merged targeted and untargeted metabolomics data revealed a significant impact of COVID-19 on amino acid metabolism, especially the pathways involving tryptophan, aspartate, arginine, tyrosine, and lysine (**Figure 2.D**). Top hits from these pathways were mapped against KEGG pathway map hsa01100 in **Figure 2.E**. Targeted absolute quantitative measurements, as determined by stable isotope-labeled internal standards, are provided in **Supplementary Table 1** for a subset of metabolites from the most significantly affected pathways.

**Figure 1.**
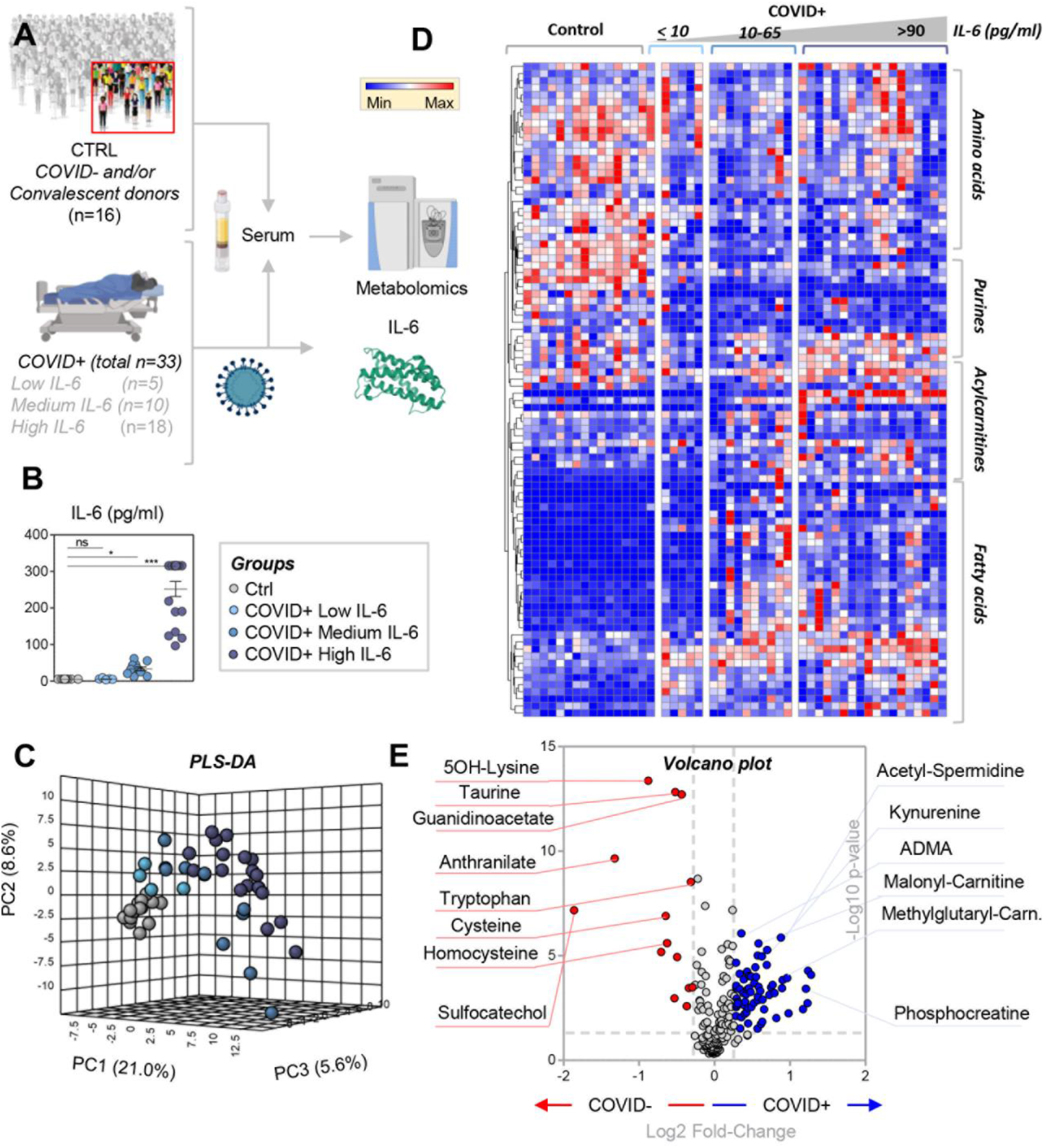
Metabolomics analysis of COVID-19 patients. Forty-nine subjects were studied, of which 16 were COVID-19-negative and 33 were COVID-19-positive patients, as determined by nucleic acid testing of nasopharyngeal swabs (**A**). IL-6 levels were determined during routine clinical care using a clinically-validated ELISA assay (**B**) and the results were used to divide COVID-19-positive patients into groups with low (<10 pg/ml), medium (10–65 pg/ml) and high (>90 pg/ml) IL-6 levels. Sera were obtained from these subjects for metabolomics analyses. In **C**, the serum metabolic phenotypes of COVID-19-positive patients significantly differed from controls by PLS-DA. Hierarchical clustering analysis (**D**) highlighted a significant impact of COVID-19 and IL-6 levels on amino acid metabolism, purines, acylcarnitines, and fatty acids. A vectorial version of this figure is provided in **Supplementary Figure 1**. The volcano plot in **E**, derived from a targeted metabolomics analysis, highlights the top serum metabolites that increase (blue) or decrease (red) in COVID-19-positive patients, as compared to controls.

**Figure 2.**
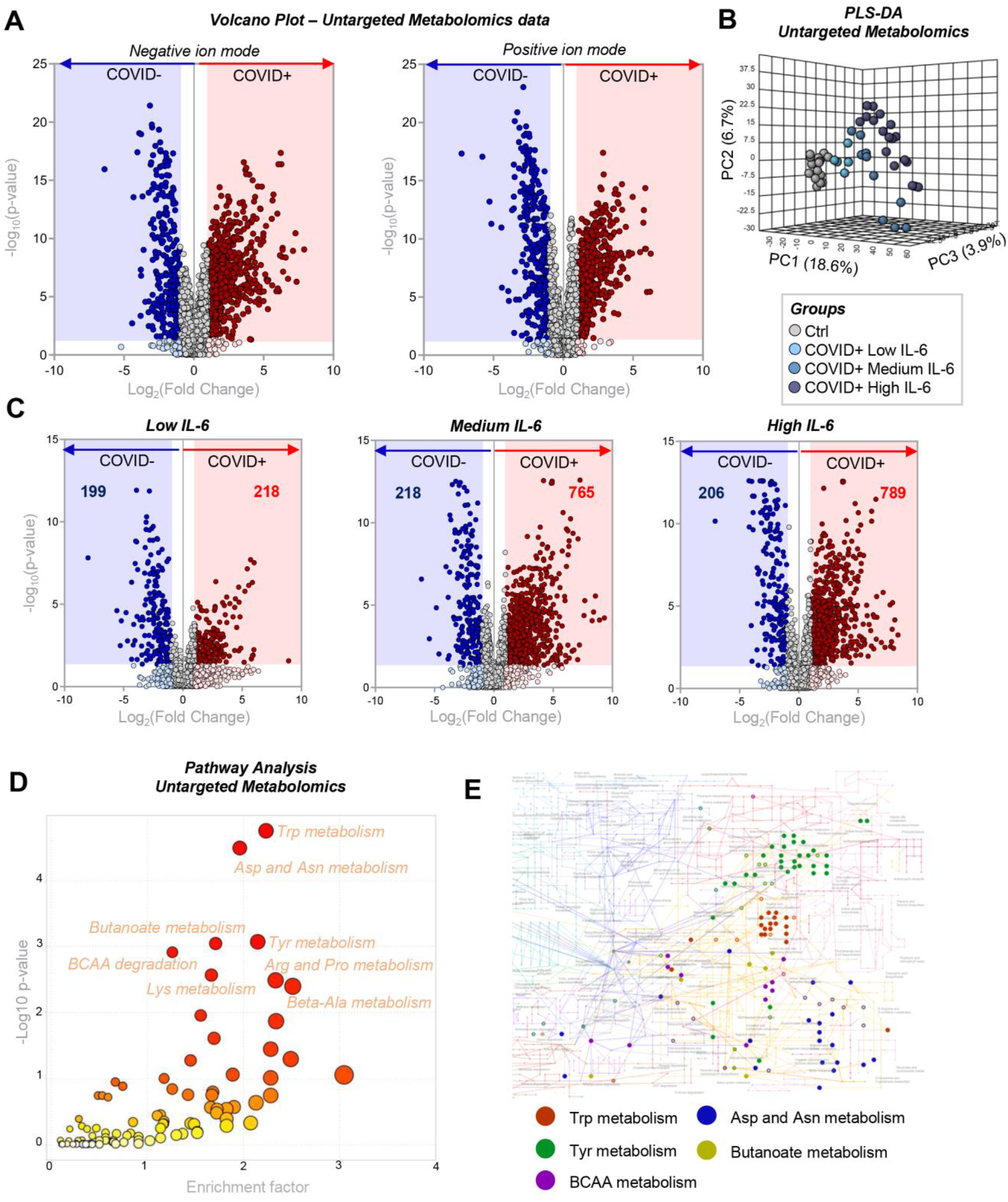
Untargeted metabolomics analyses. Untargeted metabolomics analyses were performed using sera obtained from of COVID-19-positive and –negative subjects. Volcano plots in **A** highlight 3,034 and 2,484 differential metabolites (unique molecular formulae were determined by high-resolution accurate intact mass, isotopic patterns, and MS/MS analyses) for negative and positive ion mode (left and right panel in **A**), respectively. In **B**, PLS-DA based on the untargeted metabolomics data further separates COVID-19-negative and –positive subjects, the latter separating from the former across PC1 (18.7% of the total variance) as a function of IL-6 levels. In **C**, volcano plots highlight differences between COVID-19 positive subjects with low, medium, and high serum IL-6 levels, as compared to controls. In **D**, pathway analysis of untargeted metabolomics data identified significant effects of COVID-19 on amino acids, especially regarding tryptophan, aspartate, arginine, tyrosine, and lysine metabolism. In **E**, top hits from these pathways are mapped against KEGG pathway map hsa01100.

### Alteration of tryptophan metabolism in sera of COVID-19-positive patients

Tryptophan metabolism was the top pathway affected by COVID-19 in the analysis of targeted and untargeted metabolomics data (**Figure 2.E**). As such, focused analysis of this pathway highlighted significant decreases (inversely proportional to IL-6 concentration) in tryptophan, serotonin, and indole-pyruvate levels (**Figure 3.A**). In contrast, increases in kynurenine, kynurenic acid, picolinic acid, and nicotinic acid, but not anthranilate, suggest hyperactivation of the kynurenine pathway (**Figure 3.A**). Although it is beyond the scope of this work to identify diagnostic metabolic markers of COVID-19 positivity, receiver operating characteristic (ROC) curves were calculated for the absolute quantitative measurements of serum tryptophan and kynurenine in these sera (**Figure 3.B**); interestingly, tryptophan levels lower than 105 μM and kynurenine levels higher than 5.3 μM have areas under the curves > 95% in distinguishing between the two groups (positive vs. negative); the potential relevance of this observation will require additional study in the future.

**Figure 3.**
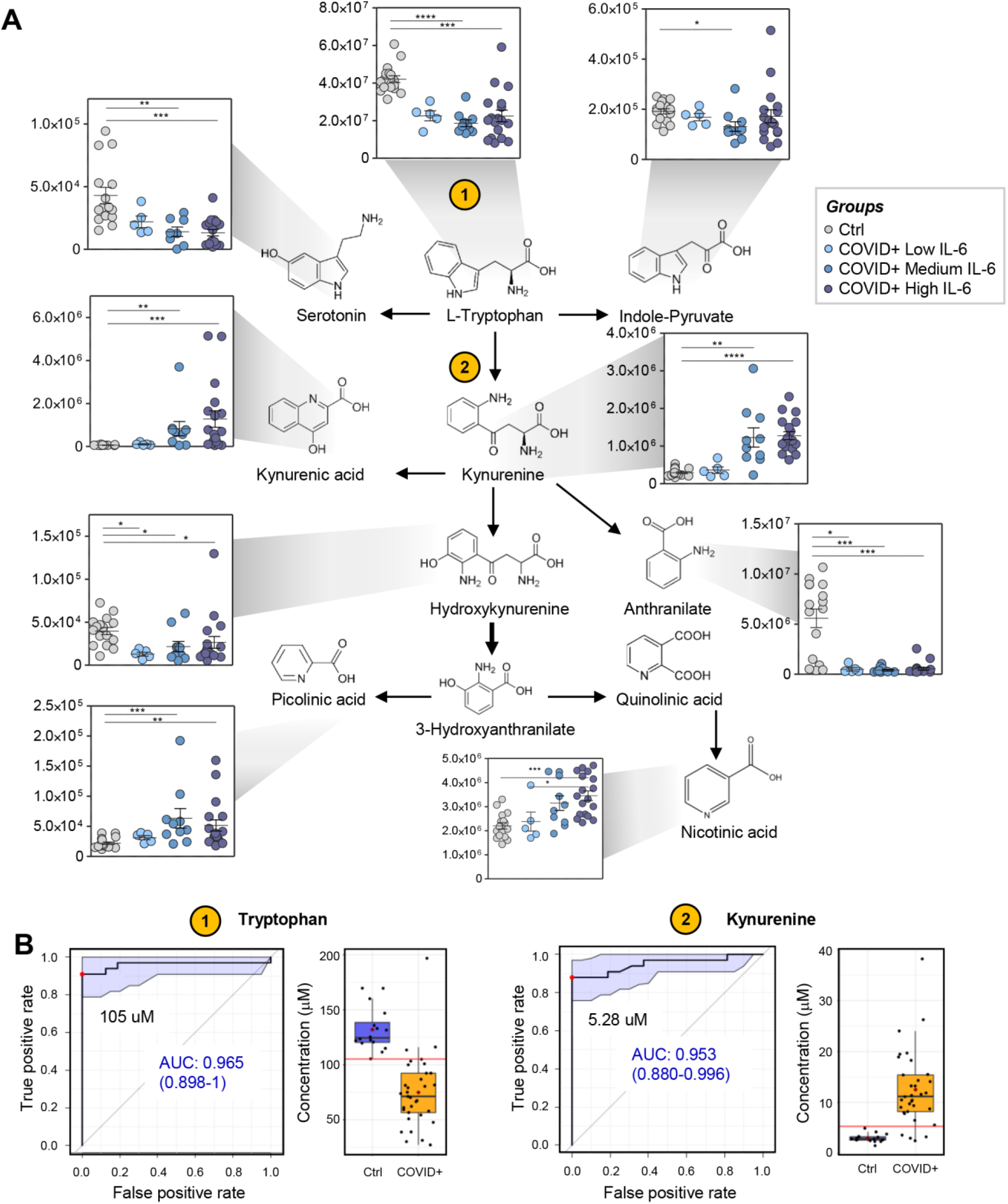
Alterations of tryptophan metabolism in COVID-19-positive subjects. Tryptophan metabolism was identified in targeted and untargeted metabolomics data as the top pathway affected by COVID-19 (**A**). Asterisks indicate significance by ANOVA (One-way ANOVA with Tukey multiple column comparisons – * *p<0.05; ** p<0.01; *** p<0.001; ****p<0.0001*). In particular, decreases in tryptophan and increases in kynurenine were proportional to disease severity, as inferred by IL-6 levels, and predicted COVID-19 infection with good sensitivity and specificity, as shown by the ROC curves in **B**.

### Dysregulation of amino acid metabolism in COVID-19 patients as a function of IL-6 levels

Beyond tryptophan, metabolomics analyses revealed a significant impact of SARS-CoV-2 infection on serum amino acid levels (**Figure 4**). Specifically, gluconeogenic and sulfur-containing amino acids (e.g., cysteine, taurine) tended to decrease, especially in the moderate-high IL-6 group (**Figure 4.A**). In contrast, oxidized forms of sulfur-containing amino acids (e.g., methionine sulfoxide, cystine), as well as of arginine, increased (**Figure 4.A**). Although no significant changes were noted in methionine levels, increases in acetyl-methionine and hydroxyproline, byproducts of proteolysis and collagen catabolism, respectively, were observed, particularly in COVID-19 patients with the highest IL-6 levels (**Figure 4.B**). Despite increases in arginine, the urea cycle metabolic intermediates ornithine and citrulline decreased in COVID-19 patients (**Figure 4.C**). In addition, creatine and creatinine, as well as the polyamines spermidine and acetyl-spermidine, increased significantly, particularly in patients with medium and high IL-6 levels (**Figure 4.C-D**); this is suggestive of renal dysfunction in these groups, which was confirmed by clinical laboratory measurements of creatinine and BUN (**Supplementary Table 1**).

**Figure 4.**
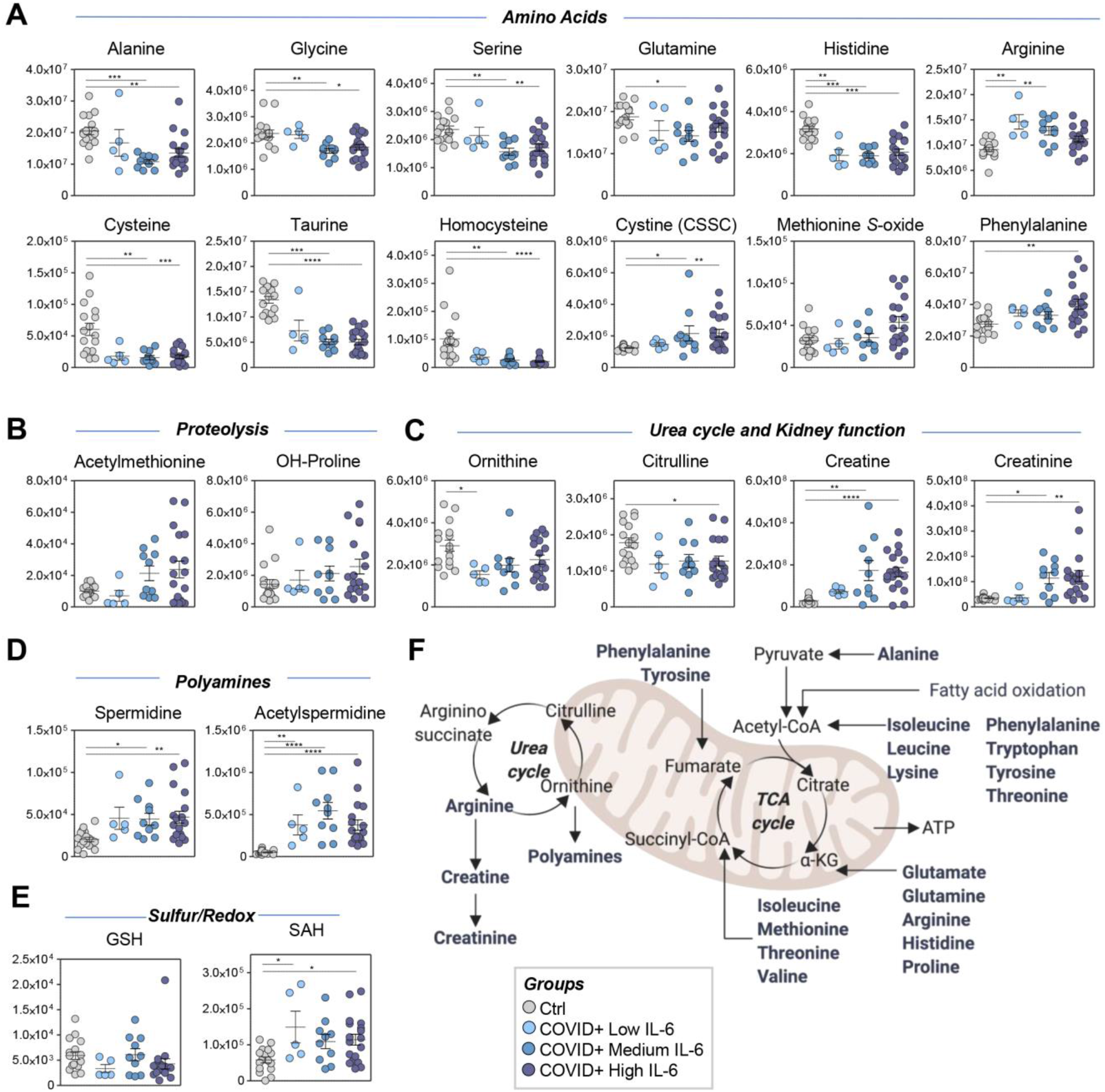
Amino acid levels and metabolism in sera of COVID-19-positive patients. Serum levels of amino acids (**A**), proteolysis markers (**B**), urea cycle and renal function-related metabolites (**C**), polyamines (**D**), and sulfur-redox metabolism (**E**) showed significant differences between COVID-19-positive patients and controls. An overview of the pathways related to these metabolites is in **F**. Asterisks indicated significance by ANOVA (One-way ANOVA with Tukey multiple column comparisons – * *p<0.05; ** p<0.01; *** p<0.001; ****p<0.0001*

### Evidence of hyperglycemia, hemolysis, and lipid abnormalities in sera of COVID-19 patients

Because altered amino acid metabolism suggested dysregulated nitrogen metabolism, we determined whether this was also accompanied by significant alterations in carbon metabolism. Notably, all COVID-19 patients in this cohort, independent of IL-6 levels, exhibited hyperglycemia (**Figure 5** – also confirmed by clinical laboratory measurements – **Supplementary Table 1**). Some metabolic intermediates of the glycolysis and pentose phosphate pathways were increased in subjects with the highest IL-6 levels; this was suggestive of potential hemolysis (e.g., ribose phosphate; **Figure 5**). However, unexpected decreases in lactate levels were noted in the same subjects, in the absence of differences in the levels of all carboxylic acids except alpha-ketoglutarate (**Figure 5**); this suggested altered transamination homeostasis consistent with an altered nitrogen balance.

**Figure 5.**
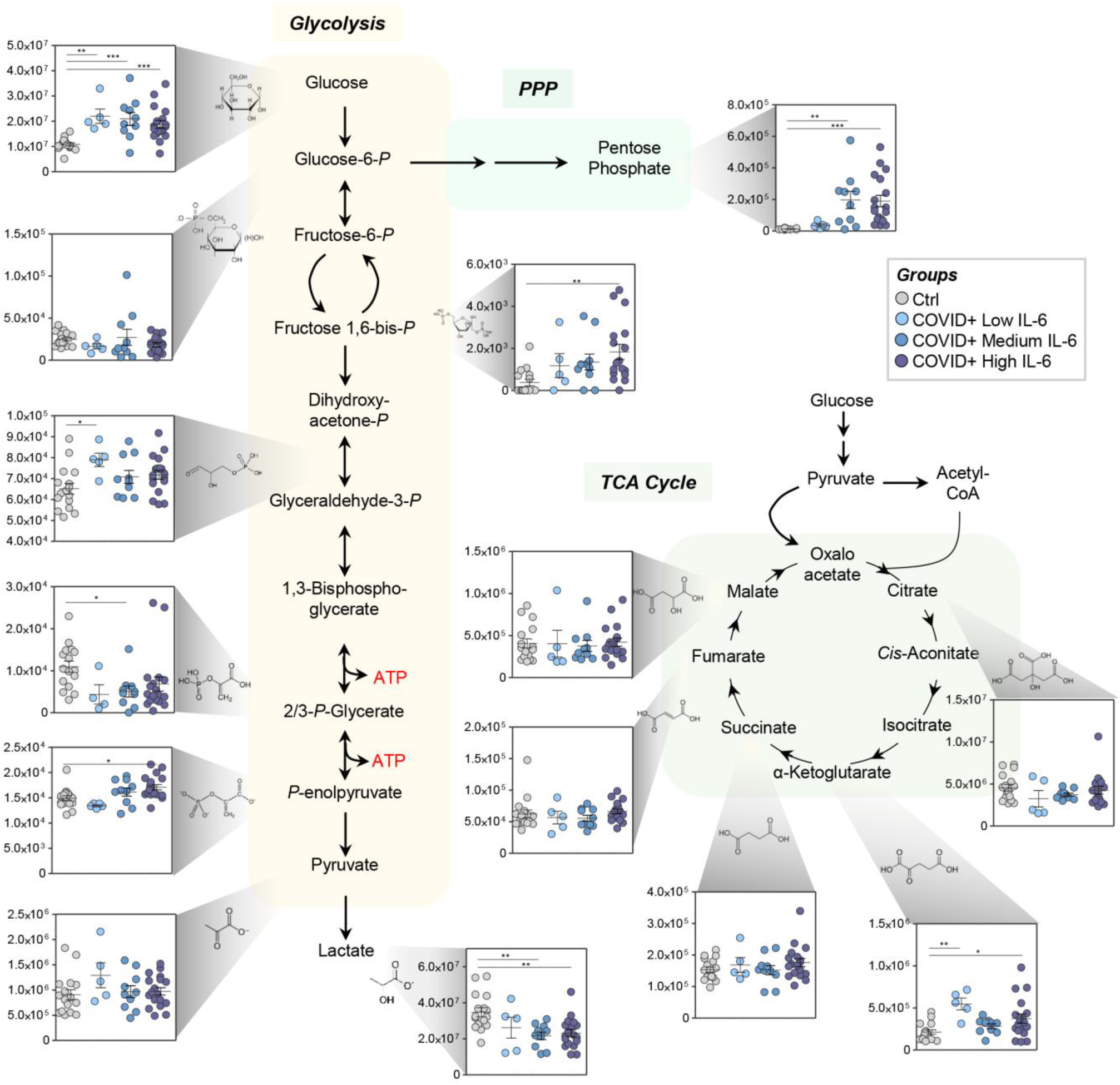
Circulating levels of glucose and its catabolites in sera of COVID-19-positive patients. Serum levels of glucose were significantly different when comparing COVID-19-positive patients and controls. Similarly, significant increases were seen in the levels of some intermediates of the glycolytic and pentose phosphate pathways in sera of COVID-19-positive patients. No significant changes were noted in serum levels of carboxylic acids across the groups. Asterisks indicated significance by ANOVA (One-way ANOVA with Tukey multiple column comparisons – * *p<0.05; ** p<0.01; *** p<0.001; ****p<0.0001*).

In addition, sera from COVID-19 patients demonstrated significant changes in levels of acylcarnitines and free fatty acids (**Figure 6**). Specifically, all short and medium-chain acylcarnitines, but not acyl-C18:3, were significantly decreased in all COVID-19 patients, independent of IL-6 levels. Finally, all fatty acids, except for nonanoic acid, were increased in all COVID-19 patients, independent of IL-6 levels (**Figure 6**).

**Figure 6.**
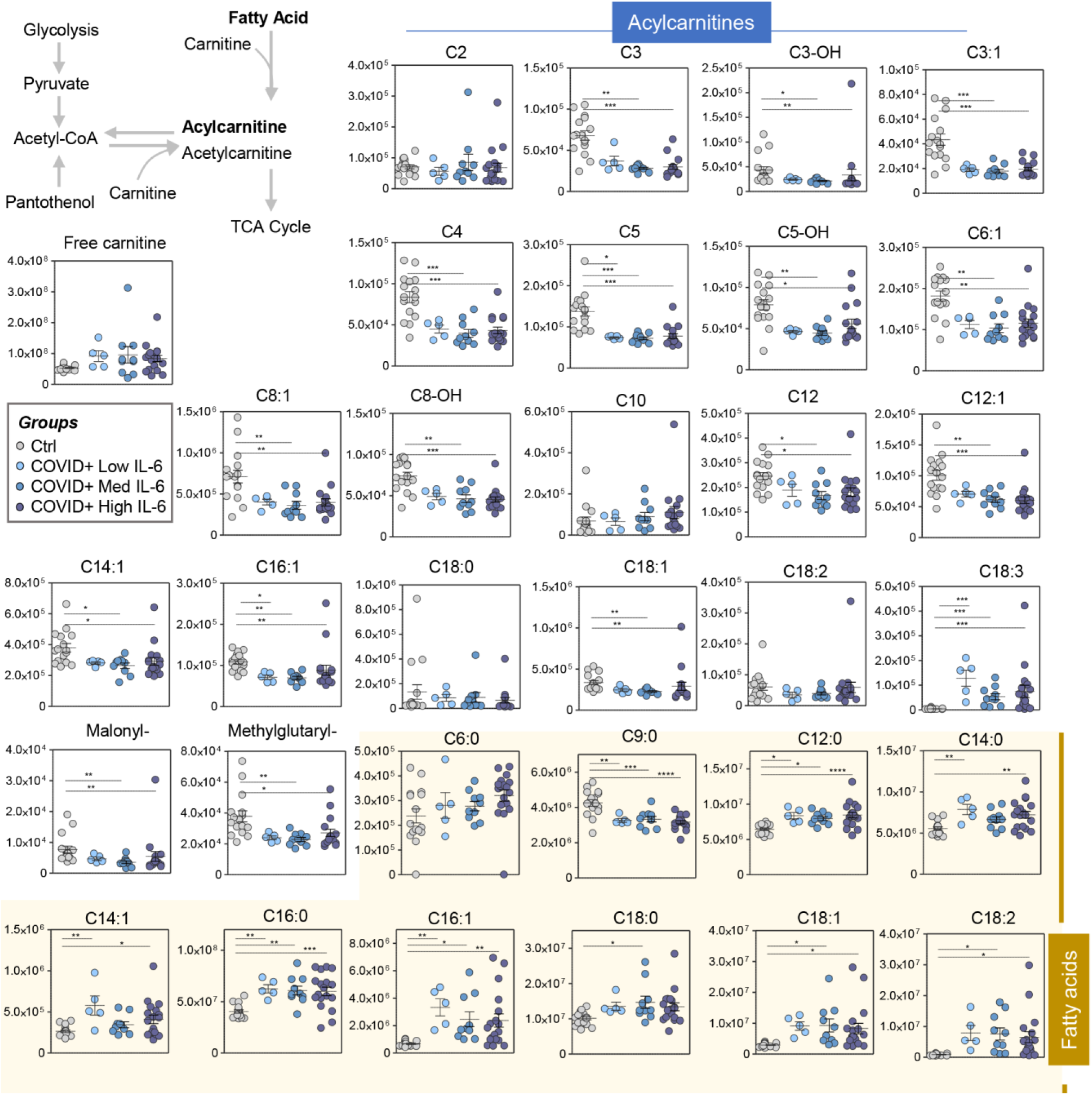
Circulating levels of free fatty acids and acylcarnitines in sera of COVID-19-positive patients. Serum levels of free fatty acids and acylcarnitines were significantly different when comparing COVID-19-positive patients and controls. Asterisks indicated significance by ANOVA (One-way ANOVA with Tukey multiple column comparisons – * *p<0.05; ** p<0.01; *** p<0.001; ****p<0.0001*).

### Metabolic correlates of laboratory markers of inflammation and renal function

To determine the potential clinical relevance of these metabolic findings, metabolite levels were compared to clinical laboratory markers of inflammation (i.e., IL-6 and C-reactive protein (CRP)) and renal function (i.e., BUN; **Figure 7.A-C**). Notably, several acylcarnitines, kynurenine, and methionine sulfoxide were among the top correlates to IL-6 (**Figure 7.A**). Interestingly, acylcarnitines, but not free carnitine, ranked among the top positive correlates to BUN levels, along with creatinine (as expected), carboxylic acids (e.g., fumarate, itaconate), and oxidized purines (e.g., inosine; **Figure 7.B**). Free fatty acid and tryptophan levels were among the top negative correlates to CRP levels, which were also directly proportional to levels of picolinic acid, inosine, and short-chain acylcarnitines (**Figure 7.C**).

**Figure 7.**
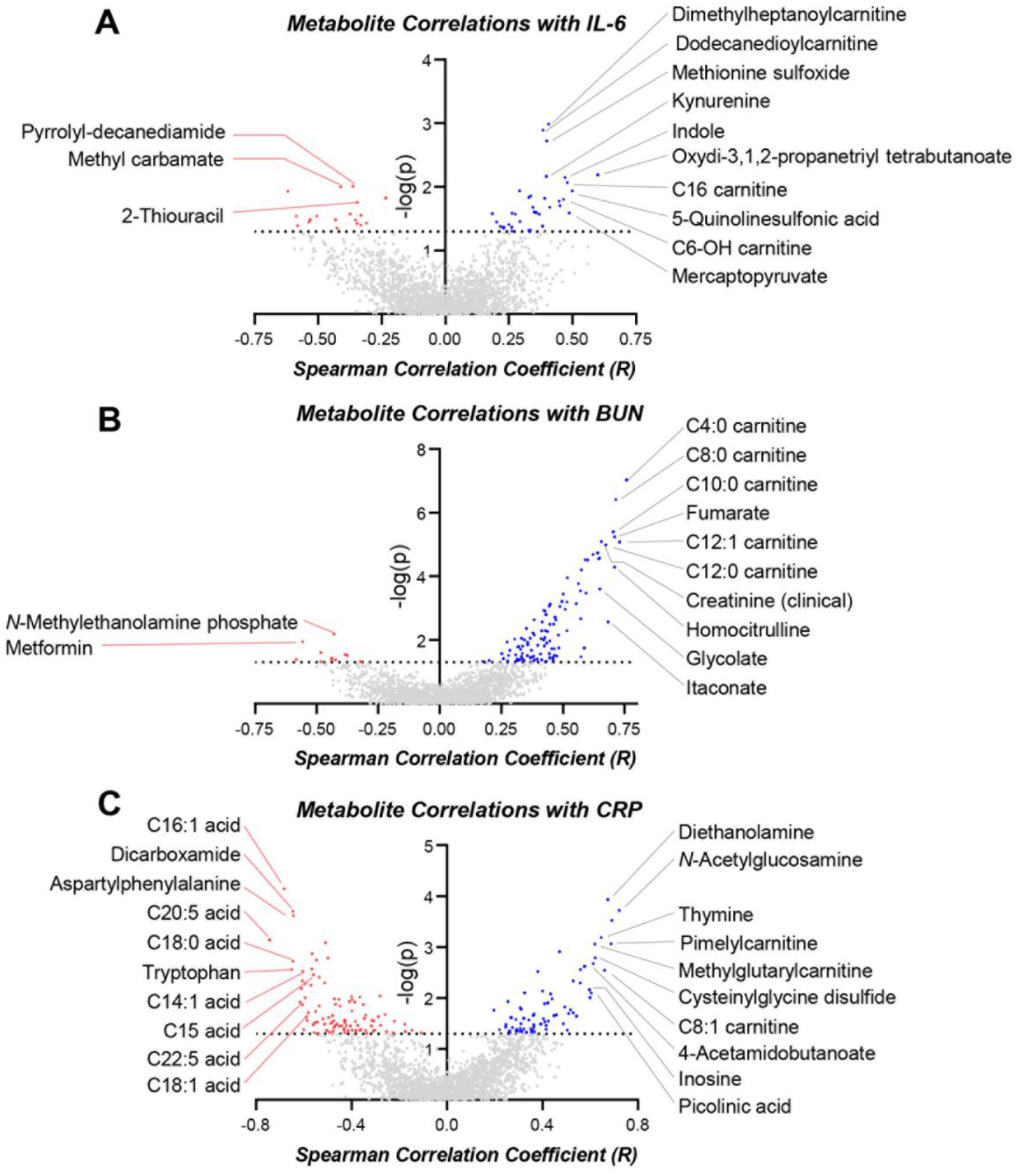
– Metabolic correlates to IL-6, renal function (BUN), and CRP. Metabolite levels were correlated (Spearman – x axis) to measurements of IL-6 (**A**), renal function (BUN; **B**) and CRP (**C**). In each panel, the xaxis indicates Spearman’s correlation coefficients and the y-axis indicates the significance of the correlation (-log10 of p-values for each correlate).

## Discussion

The present study provides the first observational metabolic characterization of sera from COVID-19-positive patients. The results show marked alterations in nitrogen metabolism (especially amino acid homeostasis, catabolism, and transamination) and carbon metabolism (especially affecting levels of glucose and free fatty acids). Metabolites in these pathways significantly correlated with circulating levels of inflammatory markers (i.e., IL-6 and CRP) and renal function (i.e., BUN). Interestingly, the major effects of COVID-19, in addition of IL-6 levels, were tryptophan metabolism and the kynurenine pathway. The rate-limiting step in this pathway is determined by the enzymatic activity of indole 2,3-dioxygenase (IDO1), a pleotropic enzyme with complex effects, including an immunoregulatory role (29). In general, the kynurenine/tryptophan ratio is a general measure of IDO activity, although the meaning of this measure alters depending upon the dynamics of metabolism and its effects on half-lives of kynurenine and tryptophan. IDO has substantial immunoregulatory effects that typically serve as a negative regulatory pathway of inflammation and immunization (30). Of particular relevance to the current study, IDO plays a significant role in limiting lung inflammation in the context of IL-6, and deletion of IDO severely exacerbated inflammatory lung pathology in a murine model (31). IDO regulates immune responses by depleting tryptophan and by generating tryptophan metabolites that activate the aryl hydrocarbon receptor (AhR); both of these metabolic alterations were observed in the current report. Moreover, IDO activity (inferred from the levels of its metabolites) is inversely correlated with IL-6 levels. Although causes and effects cannot be inferred in the current observational study, based upon known biology, it is reasonable to hypothesize that IDO activity is mechanistically involved in inflammation in COVID-19. Consistent with this hypothesis, IDO is expressed by mucosal surfaces, such as the lung, including antigen presenting cells and the epithelium (32).

IDO1is a downstream target of interferon signaling (involving the STAT6 and NF-κB pathways) (29), which is activated in response to viral infection (33). Intriguingly, however, type I interferon production is suppressed in SARS-COV-2 infection; in addition, multiple studies suggest that interferon gamma production correlates with disease severity (34–37). In one such predicted mechanism, interferon induces ACE2 receptor expression,(38) which is also the receptor for the SARS-COV-2 spike protein S that mediates infection of host target cells (10–12). Thus, interferon and its downstream signaling partners can have complex and potentially opposing effects in the context of SARS-COV-2 pathology and the ensuing balance of inflammation. Importantly, IDO activity is also regulated by oxidative stress and is enhanced by superoxide and NO. In summary, multiple factors may affect the ability of IDO to limit inflammation and affect adaptive immunity. Therefore, IDO activity may increase survival from COVID-19 by limiting pulmonary inflammation while simultaneously decreasing the development of protective adaptive immunity to SARS-COV-2.

Recently, we reported an analogous activation of the IDO pathway in individuals with Trisomy 21 (Down syndrome) (29). Indeed, 4 of the 6 receptors for interferon-alpha and –gamma are encoded by genes on chromosome 21. In addition, individuals with Down syndrome suffer from respiratory comorbidities (e.g., pulmonary hypertension, chronic obstructive pulmonary disease), gastrointestinal disorders, diabetes, and accelerated aging (29), all risk factors for, or manifestations of, severe COVID-19 illness. In the Down syndrome population, therapeutic strategies that target IDO1 or its upstream signaling pathways (i.e., involving Janus-activated kinase (JAK)), were proposed to ameliorate some comorbidities accompanying Trisomy 21 (39). In this context, the current findings suggest the possible relevance of JAK inhibitors in treating COVID-19.(40) Epidemiological studies of COVID-19 incidence and severity in the Down syndrome population could also provide clues with respect to the role of interferon signaling in response to SARS-CoV-2 infections.

Nonetheless, Blanco-Melo and colleagues reported a blunted interferon I and III response in COVID-19 (41), which could, in part, be explained by the activity of SARS-CoV-2 proteins ORF3B, ORF6, and the nsp nuclease, these were implicated in interferon antagonism and in cleaving host mRNA to prevent ribosomal loading and causing host shut-off, respectively.(42, 43) Similarly, others described impaired type I interferon activity and exacerbated inflammatory responses in severe Covid-19 patients (44), though they also described that progressive increases in disease severity, from mild to severe to critical, correlated with the levels of transcripts for JAK1, STAT1 and 2, interferon alpha 2, interferon alpha receptors 1 and 2, and interferon regulatory factors 1, 4, 5 and 7. In this view, strategies were designed to supplement interferon alpha/beta when treating COVID-19 (45), which can significantly reduce the duration of detectable virus in the upper respiratory tract and, in parallel, reduce the duration of elevated blood levels of IL-6 and CRP. Although speculative, it is interesting to note that metabolites in the kynurenine pathway are critical immunomodulators, as they contribute, along with arginine (one of the most affected pathways identified herein), to the immunosuppressive activity of dendritic cells (46) and to CD8 T cell suppression in triple negative breast cancer (47, 48). Therefore, activation of this pathway may be a mechanism by which SARS-CoV-2 promotes immune evasion. Moreover, dysregulated tryptophan metabolism is seen in inflammation (49) and aging, resulting in decreased NAD synthesis in older animals (50) and humans, an observation that supports an industry providing nicotinamide riboside as a dietary supplement (51); this suggests a potential metabolic explanation for the increased severity of COVID-19 in older subjects. In this context, it is worth noting that pro-inflammatory signaling favors proteolysis and amino acid catabolism (represented herein by increased acetyl-methionine and hydroxyproline, and altered levels of free amino acids), which can be antagonized, in part, by anti-inflammatory cytokines (e.g., IL-37)(52) or inflammasome inhibitors (53). Notably, these same anti-inflammatory cytokines show promise in regulating glycemia in aging mice (54), which is relevant in light of finding increased glycemia in our COVID-19 patients; this may also help explain why diabetes is a risk factor for severe COVID-19 illness (55). Interestingly, similar increases in circulating glucose levels are found in critically ill patients, a phenomenon termed “traumatic diabetes”.(56–58) Similarly, in trauma patients, extreme lipolysis is seen minutes to hours after trauma/shock, resulting in increased circulating free fatty acid levels (56), which was also seen herein in the context of COVID-19, especially in patients with high IL-6 levels. Indeed, a key finding in the current study was elevated long-chain free fatty acids in COVID-19 subjects. Phospholipase A2 (PLA2), which cleaves the fatty acid in the sn-2 position of phospholipids, is required for replication of other related viruses such as MERS-CoV and HCoV-229E (25). Elevated levels of long-chain polyunsaturated fatty acids (PUFA), which are highly enriched in the sn-2 position of phospholipids, suggests that this pathway may be upregulated during SARS-CoV-2 infection. Increased PLA2 activity is associated with increased production of bioactive lipids by metabolism of omega-6 PUFAs and lysophosphatidylcholine. Some of the resulting metabolites (e.g., eicosanoids, oxylipins) may help propagate infection and thrombo-inflammatory complications (59), by directly supporting synthesis of viral capsid membranes (24–26), by targeting pro-inflammatory immune cells, or by activating platelets (60). Thus, inhibiting PLA2 activity or increasing omega-3 PUFA levels, which are metabolized to bioactive lipids with anti-inflammatory activity, could potentially reduce disease severity.

Inflammatory stimuli promote platelet hyperreactivity in older individuals (22). Notably, polyamine byproducts of nitrogen metabolism (e.g., spermidine, acetyl-spermidine), were among the metabolites that increased the most COVID-19 patient sera. Primary amines, such as tranexamic acid, can prevent hyperfibrinolysis in actively bleeding patients by interacting with the Kringle domain of tissue plasminogen activator and, thereby, blocking the fibrinolytic cascade (61). It is interesting to speculate that inflammation-induced alterations in nitrogen metabolism may participate in dysregulated coagulation leading to thrombus formation in the most severe cases of COVID-19. In contrast, other metabolites associated with hypercoagulability, such as homocysteine (62) (which increases in Down syndrome (63)), were decreased in COVID-19 patient sera. The latter may be explained by the apparent increase in oxidant stress in COVID-19 patients, as suggested by increased serum levels of methionine sulfoxide and cystine, along with decreases in antioxidants, such as cysteine and taurine (64) (but, interestingly, not reduced glutathione). Conversely, decreased serum acylcarnitine levels, which may have anticoagulant function (65), is consistent with a potential hypercoagulable phenotype in these patients.

Interestingly, increased serum sphingosine 1-phosphate levels were observed in COVID-19 patients. Circulating levels of this metabolite are significantly influenced by red blood cell (RBC) Sphingosine Kinase 1 activity in response to hypoxia (66), whereas alterations to this pathway mediate responses to angiotensin II-induced stimulation in the hypoxic kidney in chronic kidney disease (67); the latter may be relevant in light of renal dysfunction in some of the COVID-19 patients in the current study. In contrast, one would have predicted significant increases in serum levels of lactate and carboxylic acid markers of hypoxia (e.g., succinate) in COVID-19 patients, similar to what is seen in hypoxemic patients following ischemic (68) or hemorrhagic shock (69). Surprisingly, no significant increases in serum lactate or succinate levels were observed, which may be explained because our patients were either normoxic or receiving intense respiratory therapy (e.g., ventilation) at the time of the blood draw. Although unrelated to IL-6, fumarate and itaconate were positively correlated with BUN and inosine, suggesting potential cross-talk between deaminated purine salvage reactions and carboxylic acid metabolism in renal dysfunction in COVID-19, similar to acute kidney ischemia (70). In addition, hemolysis markers (e.g., metabolic intermediates of the glycolytic and pentose phosphate pathways) increased proportionally to IL-6 levels, suggesting a correlation between inflammation, disease severity, and, ultimately, the extent of therapeutic interventions and related mechanical/oxidant stress on circulating RBCs. As such, future studies will be designed to assess COVID-19 effects on RBCs.

The present study has several limitations. First, the analyses were performed on sera obtained as a byproduct of routine clinical laboratory testing. Although immediately refrigerated and stored overnight for less than 24h, technical bias may have been introduced and propagated across all samples owing to the collection procedures. Protocols are currently being implemented to prospectively collect and bank fresh samples of serum and plasma, red blood cells, and buffy coats. In addition, males are disproportionally represented in our study: 75% of the COVID-19-positive patients. As such, no sex-specific analysis was performed, and future investigations on larger, prospectively enrolled cohorts are currently underway to address this issue. Similarly, the mean age of COVID-19-positive patients was 56, whereas the control population was generally younger (mean age: 33 years old). Nonetheless, comparisons to prior studies on the effect of aging on the plasma/whole blood metabolome does not suggest that the observations reported here are due to age alone (71, 72). In addition, the COVID-19 patients in this study had significant disease, by definition, because they were all inpatients; future studies will investigate patients with milder disease and asymptomatic infected patients. Finally, it will be important to study serial samples from patients throughout their clinical course to evaluate changes in the serum metabolome as a function of clinical status (e.g., on or off a ventilator, receiving hemodialysis or not) and in response to various therapeutic interventions (e.g., treatment with IL-6 inhibitors).

In conclusion, herein we provide the first observational report of the comprehensive serum metabolome in COVID-19-positive patients as a function of IL-6 level, the latter as a proxy for disease severity and as a promising therapeutic target. Our results suggest several potential therapeutic targets in systems metabolism, potentially through dietary and pharmacological intervention. These include (i) IDO1 or its upstream modulators (e.g., JAK, STAT, and/or interferon signaling, although we cannot predict whether inhibition or further stimulation of this pathway would be beneficial for COVID-19 pathology and/or immunity); (ii) amino acid catabolism as a function of oxidant stress (e.g., taurine) or inflammation-induced proteolysis (targetable directly through dietary supplementation with taurine/N-acetylcysteine, or indirectly by using antibodies against pro-inflammatory cytokines or antagonists to their receptors (e.g., anakinra), or by administering anti-inflammatory cytokines or inflammasome inhibitors (52–54)); and (iii) free fatty acid levels and unsaturation, which are amenable to dietary or pharmacological intervention (e.g., fish oil diet or fatty acid desaturase inhibitors).

## Methods

### Blood collection and processing

This observational study was conducted according to the Declaration of Helsinki, in accordance with good clinical practice guidelines, and approved by the Columbia University Institutional Review Board. Subjects seen at Columbia University Irving Medical Center/New York-Presbyterian Hospital included 33 COVID-19-positive patients, as determined by SARS-CoV-2 nucleic acid testing of nasopharyngeal swabs; in this group, the severity of the disease was inferred from serum IL-6 levels, which were determined by a CLIA-certified ELISA-based assay. The control group included 16 subjects, all of whom were SARS-CoV-2 negative by nasopharyngeal swab at the time of the blood draw. Some patients in this group were “never positive” subjects and some were COVID-19 convalescent patients who were previously positive, but currently negative, as determined by testing nasopharyngeal swabs, and at least 14 days post-resolution of symptoms. Serum was obtained from freshly drawn blood after an overnight hold at 4ºC and all samples were then de-identified. Sera were extracted via a modified Folch method (chloroform/methanol/water 8:4:3), which completely inactivates other coronaviruses, such as MERS-CoV (73). Briefly, 20 μL of serum was diluted in 130 μl of LC-MS grade water, 600 μl of ice-cold chloroform/methanol (2:1) was added, and the samples vortexed for 10 seconds. Samples were then incubated at 4ºC for 5 minutes, quickly vortexed (5 seconds), and centrifuged at 14,000 x *g* for 10 minutes at 4ºC. The top (i.e., aqueous) phase was transferred to a new tube for metabolomics analysis.

### Ultra-High-Pressure Liquid Chromatography-Mass Spectrometry (MS) metabolomics

Metabolomics analyses were performed using a Vanquish UHPLC coupled online to a Q Exactive mass spectrometer (ThermoFisher, Bremen, Germany). Samples were analyzed using 5, 15, and 17 min gradients, as described (74, 75). For targeted quantitative experiments, extraction solutions were supplemented with stable isotope-labeled standards, and endogenous metabolite concentrations were quantified against the areas calculated for heavy isotopologues for each internal standard (74, 75). Data were analyzed using Maven (Princeton University) and Compound Discoverer 2.1 (ThermoFisher). Graphs and statistical analyses were prepared with GraphPad Prism 8.0 (GraphPad Software, Inc, La Jolla, CA), GENE E (Broad Institute, Cambridge, MA, USA), and MetaboAnalyst 4.0.(76) Spearman’s correlations and related p-values were calculated with R Studio.

## Data Availability

All raw data and statistical elaborations are provided as supplementary files

## Acknowledgments

This research was supported by funds from the Boettcher Webb-Waring Investigator Award (ADA), RM1GM131968 (ADA and KCH) from the National Institute of General and Medical Sciences, and R01HL146442 (ADA), R01HL149714 (ADA), R01HL148151 (ADA, SLS), R21HL150032 (ADA), and T32 HL007171 (TN) from the National Heart, Lung, and Blood Institute.

## Authors’ contributions

TT, ROF, SLS, EAH designed the study. TT, ROF, EAH collected and processed the samples. DS, LB, JAR, TN, ADA performed metabolomics analyses and prepared the figures. JAR, TN, and ADA performed data analysis and prepared figures and tables. ADA wrote the first draft of the manuscript, which was significantly revised by SLS, TT, JCZ and KEH and finally approved by all the authors.

## Disclosure of Conflict of interest

Though unrelated to the contents of this manuscript, the authors declare that AD, KCH, and TN are founders of Omix Technologies Inc and Altis Biosciences LLC. AD and SLS are consultants for Hemanext Inc. SLS is also a consultant for Tioma, Inc. JCZ is a consultant for Rubius Therapeutics. All the other authors disclose no conflicts of interest relevant to this study.

